# Estimating the wave 1 and wave 2 infection fatality rates from SARS-CoV-2 in India

**DOI:** 10.1101/2021.05.25.21257823

**Authors:** Soumik Purkayastha, Ritoban Kundu, Ritwik Bhaduri, Daniel Barker, Michael Kleinsasser, Debashree Ray, Bhramar Mukherjee

## Abstract

**Objective:** There has been much discussion and debate around the underreporting of COVID-19 infections and deaths in India. In this short report we first estimate the underreporting factor for infections from publicly available data released by the Indian Council of Medical Research on reported number of cases and national seroprevalence surveys. We then use a compartmental epidemiologic model to estimate the undetected number of infections and deaths, yielding estimates of the corresponding underreporting factors. We compare the serosurvey based ad hoc estimate of the infection fatality rate (IFR) with the model-based estimate. Since the first and second waves in India are intrinsically different in nature, we carry out this exercise in two periods: the first wave (April 1, 2020 - January 31, 2021) and part of the second wave (February 1, 2021 – May 15, 2021). The latest national seroprevalence estimate is from January 2021, and thus only relevant to our wave 1 calculations.

**Results:** Both wave 1 and wave 2 estimates qualitatively show that there is a large degree of “covert infections” in India, with model-based estimated underreporting factor for infections as ***11*.*11 (95% credible interval (CrI) 10*.*71-11*.*47)*** and for deaths as ***3*.*56 (95% CrI 3*.*48 - 3*.*64)*** for wave 1. For wave 2, underreporting factor for infections escalate to ***26*.*77 (95% CrI 24*.*26 – 28*.*81)*** and to ***5*.*77 (95% CrI 5*.*34 – 6*.*15)*** for deaths. If we rely on only reported deaths, the IFR estimate is *0*.*13%* for wave 1 and ***0*.*03%*** for part of wave 2. Taking underreporting of deaths into account, the IFR estimate is ***0*.*46%*** for wave 1 and ***0*.*18%*** for wave 2 (till May 15). Combining waves 1 and 2, as of May 15, while India reported a total of nearly 25 million cases and 270 thousand deaths, the estimated number of infections and deaths stand at 491 million (36% of the population) and 1.21 million respectively, yielding an estimated (combined) infection fatality rate of ***0*.*25%***. There is considerable variation in these estimates across Indian states. Up to date seroprevalence studies and mortality data are needed to validate these model-based estimates.

## Introduction

In late August 2020, India was predicted to surpass the United States in terms of reported case counts from SARS-CoV-2 infections. To the surprise of many modelers the curve turned corner in late September with the highest number (97,894) of daily new cases reported on 16 September 2020^1^. After a steady decline for nearly five months, the curve started rising again, growing into an astronomic second wave. The highest number (414,280) of daily new cases in wave 2 was reported on May 6, 2021. As of May 15, 2021, India has reported 24.7 million cases, the second highest in the world, and nearly 270 thousand deaths, the third highest in the world. In this brief report, we reconcile estimates of the infection fatality rate (IFR) inferred from seroprevalence studies with epidemiologic model-based estimates that account for underreporting of infections and deaths in India for wave 1. We then proceed to compute, compare and combine wave 1 with wave 2 IFR estimates.

## Methods

### Synthesizing evidence from seroprevalence studies

We review available seroprevalence results that vary across states and specifically across rural versus urban areas. Whereas in many major metros and slum areas the seroprevalences were reported to be more than 50%, in rural areas there is a wide variation ***(Table 1)***. The latest national serosurvey (from 17 December 2020 to 8 January 2021) reports 21.4% of all Indians above age 18 have antibodies present that indicate past SARS-CoV-2 infection^2^. Since approximately 59%^3^ of India’s 1.36 billion citizens are above age 18 and 10.45 million infections were reported as of 8 January 2021, this points to approximately 172.47 million infections, with an implied underreporting factor of ***16*.*5 (172*.*47/10*.*45)***. In other words, only 6% of India’s COVID-19 infections are reported, while 94% remained undetected or unreported. We use this estimated number of infections to calculate the IFR. Regional studies based on crematorium data and counting obituaries in India have suggested an underreporting factor in the range of 2 to 5 for COVID-deaths; this is at best ad hoc and anecdotal in nature and no rigorous quantification of missing death numbers is currently available^4^.

**Table 1:**
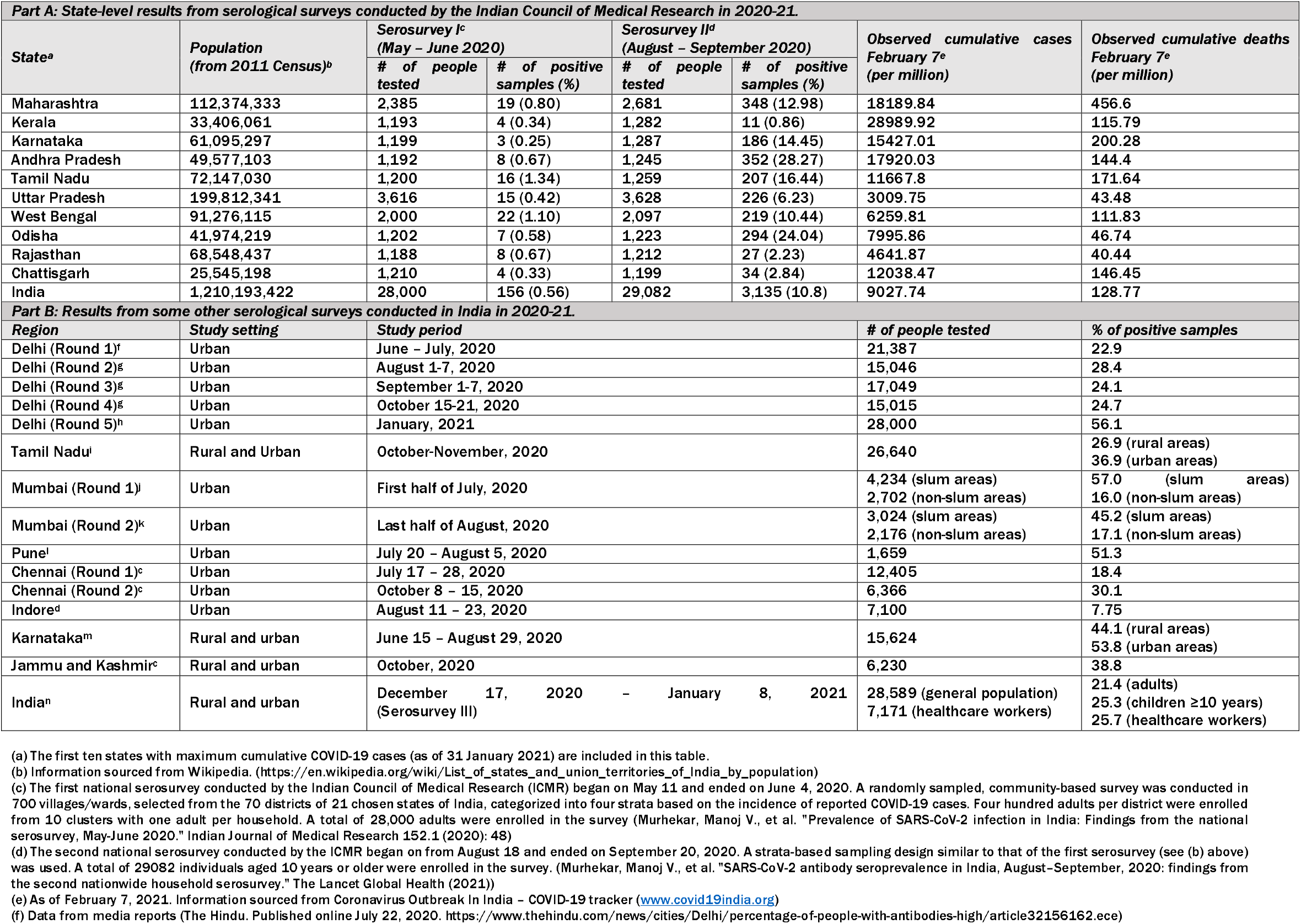
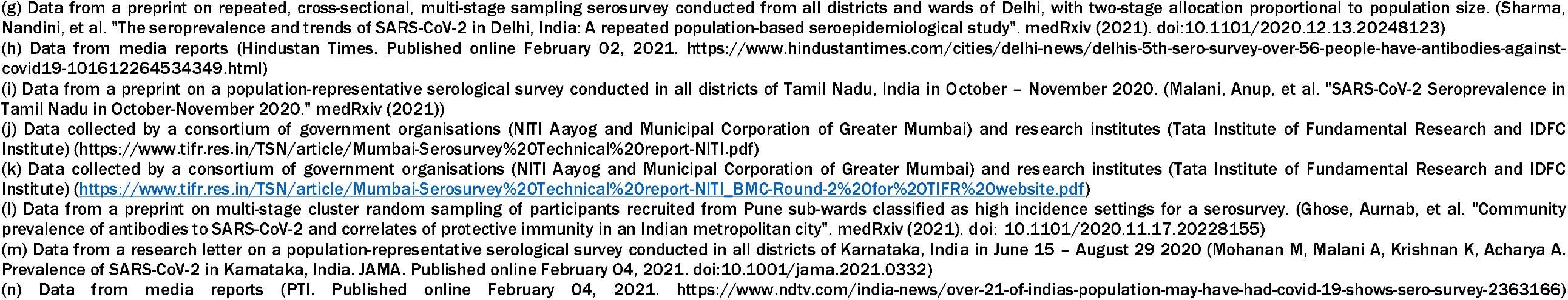
Summary of results from various serological surveys conducted in India during 2020-21.

| **Part A: State-level results from serological surveys conducted by the Indian Council of Medical Research in 2020-21** |  |  |  |  |  |  |  |
| --- | --- | --- | --- | --- | --- | --- | --- |
| **State<sup>a</sup>** | **Population**<br>**(from 2011 Census)<sup>b</sup>** | **Serosurvey I<sup>c</sup>**<br>**(May – June 2020)** |  | **Serosurvey II<sup>d</sup>**<br>**(August – September 2020)** |  | **Observed cumulative cases**<br>**February 7<sup>e</sup>**<br>**(per million)** | **Observed cumulative deaths**<br>**February 7<sup>e</sup>**<br>**(per million)** |
|  |  | **# of people**<br>**tested** | **# of positive**<br>**samples (%)** | **# of people**<br>**tested** | **# of positive**<br>**samples (%)** |  |  |
| Maharashtra | 112,374,333 | 2,385 | 19 (0.80) | 2,681 | 348 (12.98) | 18189.84 | 456.6 |
| Kerala | 33,406,061 | 1,193 | 4 (0.34) | 1,282 | 11 (0.86) | 28989.92 | 115.79 |
| Karnataka | 61,095,297 | 1,199 | 3 (0.25) | 1,287 | 186 (14.45) | 15427.01 | 200.28 |
| Andhra Pradesh | 49,577,103 | 1,192 | 8 (0.67) | 1,245 | 352 (28.27) | 17920.03 | 144.4 |
| Tamil Nadu | 72,147,030 | 1,200 | 16 (1.34) | 1,259 | 207 (16.44) | 11667.8 | 171.64 |
| Uttar Pradesh | 199,812,341 | 3,616 | 15 (0.42) | 3,628 | 226 (6.23) | 3009.75 | 43.48 |
| West Bengal | 91,276,115 | 2,000 | 22 (1.10) | 2,097 | 219 (10.44) | 6259.81 | 111.83 |
| Odisha | 41,974,219 | 1,202 | 7 (0.58) | 1,223 | 294 (24.04) | 7995.86 | 46.74 |
| Rajasthan | 68,548,437 | 1,188 | 8 (0.67) | 1,212 | 27 (2.23) | 4641.87 | 40.44 |
| Chhattisgarh | 25,545,198 | 1,210 | 4 (0.33) | 1,199 | 34 (2.84) | 12038.47 | 146.45 |
| India | 1,210,193,422 | 28,000 | 156 (0.56) | 29,082 | 3,135 (10.8) | 9027.74 | 128.77 |
| **Part B: Results from some other serological surveys conducted in India in 2020-21** |  |  |  |  |  |  |  |
| **Region** | **Study setting** | **Study period** |  | **# of people tested** |  | **% of positive samples** |  |
| Delhi (Round 1)<sup>f</sup> | Urban | June – July, 2020 |  | 21,387 |  | 22.9 |  |
| Delhi (Round 2)<sup>g</sup> | Urban | August 1-7, 2020 |  | 15,046 |  | 28.4 |  |
| Delhi (Round 3)<sup>g</sup> | Urban | September 1-7, 2020 |  | 17,049 |  | 24.1 |  |
| Delhi (Round 4)<sup>g</sup> | Urban | October 15-21, 2020 |  | 15,015 |  | 24.7 |  |
| Delhi (Round 5)<sup>h</sup> | Urban | January, 2021 |  | 28,000 |  | 56.1 |  |
| Tamil Nadu<sup>i</sup> | Rural and Urban | October-November, 2020 |  | 26,640 |  | 26.9 (rural areas)<br>36.9 (urban areas) |  |
| Mumbai (Round 1)<sup>j</sup> | Urban | First half of July, 2020 |  | 4,234 (slum areas)<br>2,702 (non-slum areas) |  | 57.0 (slum areas)<br>16.0 (non-slum areas) |  |
| Mumbai (Round 2)<sup>k</sup> | Urban | Last half of August, 2020 |  | 3,024 (slum areas)<br>2,176 (non-slum areas) |  | 45.2 (slum areas)<br>17.1 (non-slum areas) |  |
| Pune<sup>l</sup> | Urban | July 20 – August 5, 2020 |  | 1,659 |  | 51.3 |  |
| Chennai (Round 1)<sup>c</sup> | Urban | July 17 – 28, 2020 |  | 12,405 |  | 18.4 |  |
| Chennai (Round 2)<sup>c</sup> | Urban | October 8 – 15, 2020 |  | 6,366 |  | 30.1 |  |
| Indore<sup>d</sup> | Urban | August 11 – 23, 2020 |  | 7,100 |  | 7.75 |  |
| Karnataka<sup>m</sup> | Rural and urban | June 15 – August 29, 2020 |  | 15,624 |  | 44.1 (rural areas)<br>53.8 (urban areas) |  |
| Jammu and Kashmir<sup>c</sup> | Rural and urban | October, 2020 |  | 6,230 |  | 38.8 |  |
| India<sup>n</sup> | Rural and urban | December 17, 2020 – January 8, 2021<br>(Serosurvey III) |  | 28,589 (general population)<br>7,171 (healthcare workers) |  | 21.4 (adults)<br>25.3 (children ≥10 years)<br>25.7 (healthcare workers) |  |

(a) The first ten states with maximum cumulative COVID-19 cases (as of 31 January 2021) are included in this table.
(b) Information sourced from Wikipedia. (https://en.wikipedia.org/wiki/List_of_states_and_union_territories_of_India_by_population)
(c) The first national serosurvey conducted by the Indian Council of Medical Research (ICMR) began on May 11 and ended on June 4, 2020. A randomly sampled, community-based survey was conducted in 700 villages/wards, selected from the 70 districts of 21 chosen states of India, categorized into four strata based on the incidence of reported COVID-19 cases. Four hundred adults per district were enrolled from 10 clusters with one adult per household. A total of 28,000 adults were enrolled in the survey (Murhekar, Manoj V., et al. "Prevalence of SARS-CoV-2 infection in India: Findings from the national serosurvey, May-June 2020." Indian Journal of Medical Research 152.1 (2020): 48)
(d) The second national serosurvey conducted by the ICMR began on from August 18 and ended on September 20, 2020. A strata-based sampling design similar to that of the first serosurvey (see (b) above) was used. A total of 29082 individuals aged 10 years or older were enrolled in the survey. (Murhekar, Manoj V., et al. "SARS-CoV-2 antibody seroprevalence in India, August–September, 2020: findings from the second nationwide household serosurvey." The Lancet Global Health (2021))
(e) As of February 7, 2021. Information sourced from Coronavirus Outbreak In India – COVID-19 tracker (www.covid19india.org)
(f) Data from media reports (The Hindu. Published online July 22, 2020. https://www.thehindu.com/news/cities/Delhi/percentage-of-people-with-antibodies-high/article32156162.ece)

### Model-based estimates

Using a compartmental epidemiologic model (as explained in the Supplementary Methods) with a compartment for unascertained cases and deaths after accounting for the false negative rates of RT-PCR and rapid antigen tests used in India^5^ we estimate the national and state-level IFR in India by inferring underreporting factors for cases and deaths. We assume that the estimated total infections (deaths) are comprised of reported and unreported infections (deaths). The model divides the population into ten disjoint compartments: S (Susceptible), E (Exposed), T (Tested), U (Untested), P (Tested positive), F (Tested False Negative), RR (Reported Recovered), RU (Unreported Recovered), DR (Reported Deaths) and DU (Unreported Deaths), as described in ***Supplementary Figure S1***. A set of nine differential equations govern the transmission dynamics, which are approximated by means of discrete recurrence relations. For any compartment *X*, the instantaneous rate of change at time *t* (given by 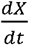) is approximated by the difference of counts in that specific compartment on the (*t* +1) th day and the (*t*) th day, i.e., say *X* (*t* + 1) *– X*(*t*). Parameters are estimated using Bayesian techniques by generating samples from the posterior distribution using a Metropolis-Hastings algorithm with Gaussian proposal density, with 95% credible intervals (CrI) to quantify uncertainty of the estimates. ***Supplementary Table T4*** presents an overview of the parameter descriptions and settings for this model.

### Comparing and combining waves 1 and 2

Due to the stress on the healthcare and reporting infrastructure, the fatality and underreporting processes were very different across the two waves. Thus, we consider two separate phases of the pandemic, with wave 1 from April 1, 2020 - January 31, 2021 and wave 2 starting on February 1, 2021. This definition is artificial and is guided by the fact that the national effective reproduction number (R_eff_) crossed unity for the first time in 2021 on February 14 and we allow a two-week incubation period before that date. Using daily time series of case, death and recovery counts we compare fatality rates and underreporting factors associated with the two time periods using the compartmental models. Further, using observed data from the two waves and the model-based underreporting factor estimates, we compute cumulative case and death counts for the total duration of waves 1 and 2. We multiply the wave-specific cumulative counts with relevant underreporting factors and sum over both waves to get combined counts of cases and deaths. The estimated numbers of cumulative deaths and infections provide us with a combined IFR estimate for India as of May 15.

Results

### IFR estimates for wave 1 using seroprevalence surveys

The observed case fatality rate (CFR) in India is low. With 154,428 deaths and 10.76 million cases reported as of January 31, 2021 the estimated CFR for wave 1 is ***1*.*435% (95% confidence interval 1*.*428% 1*.*442%)***^***1***^. The estimated number of infections from the January seroprevalence survey imply an approximate infection fatality rate of ***0*.*09%*** (i.e. 154,428/172.47M). The anecdotal underreporting factor for deaths (in the range of 2-5) implies an ad hoc estimate of IFR in the range of ***0*.*19% 0*.*45%***.

### Estimates from epidemiological models

For wave 1 our estimate for the national IFR_1_ (observed cumulative deaths/estimated cumulative total infections) is ***0*.*129% (95% CrI 0*.*125% 0*.*134%)*** and IFR_2_ (estimated total cumulative deaths/estimated total cumulative infections) is ***0*.*461% (95% CrI 0*.*455% 0*.*468%)*** with an underreporting factor for cases estimated at ***11*.*11 (95% CrI 10*.*71 11*.*47)*** and for deaths at ***3*.*56 (95% CrI 3*.*48 – 3*.*64)***. These model-based estimates in wave 1 are largely consistent with the estimates from the latest and third nationwide seroprevalence study.

In wave 2, using the same model we see a stark contrast with wave 1, with case and death underreporting factor estimates escalate to ***26*.*73 (95% CrI 24*.*26-28*.*81)*** and 5.77 ***(95% CrI 5*.*34-6*.*15)*** respectively, leading to IFR_1_ estimate of ***0*.*032% (95% CrI 0*.*029%-0*.*035%)*** and IFR_2_ estimate of ***0*.*183% (95% CrI 0*.*18%-0*.*186%)***. This pattern is consistent with wave 2 CFR being estimated at ***0*.*845% (95% CrI 0*.*840%-0*.*849%), 59%*** of wave 1 estimate.

***Figure 1*** shows underreporting factors and estimated infections and deaths in waves 1 and 2 for India while ***Figure 2*** highlights state-level variations in IFR_1_, IFR_2_, CFR for waves 1 and 2 for 20 states in India with large case/death counts.

**Figure 1 Legend:**
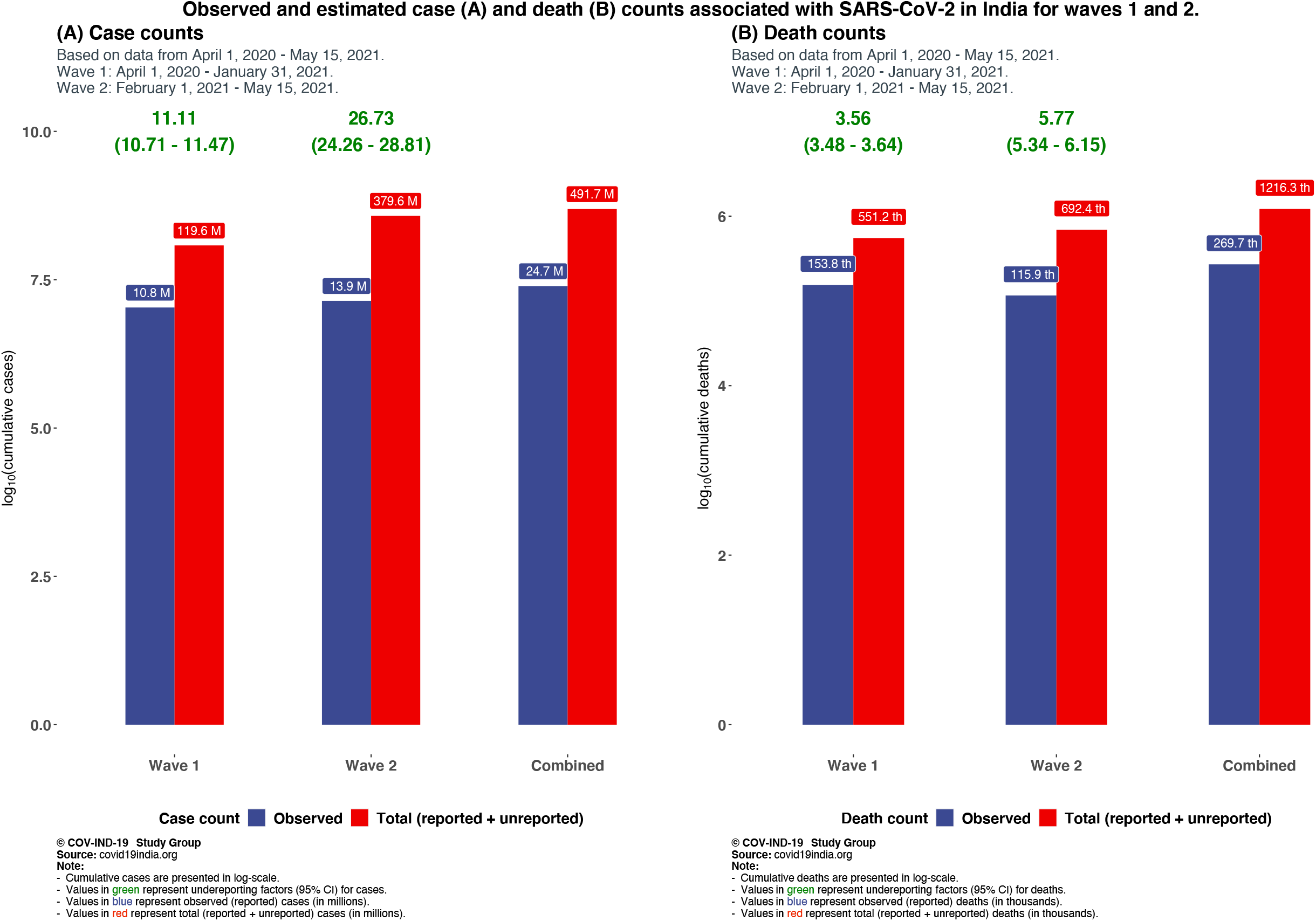
Comparison of observed and estimated case and death counts and associated underreporting factors from waves 1, 2 and both waves combined.

**Figure 2 Legend:**
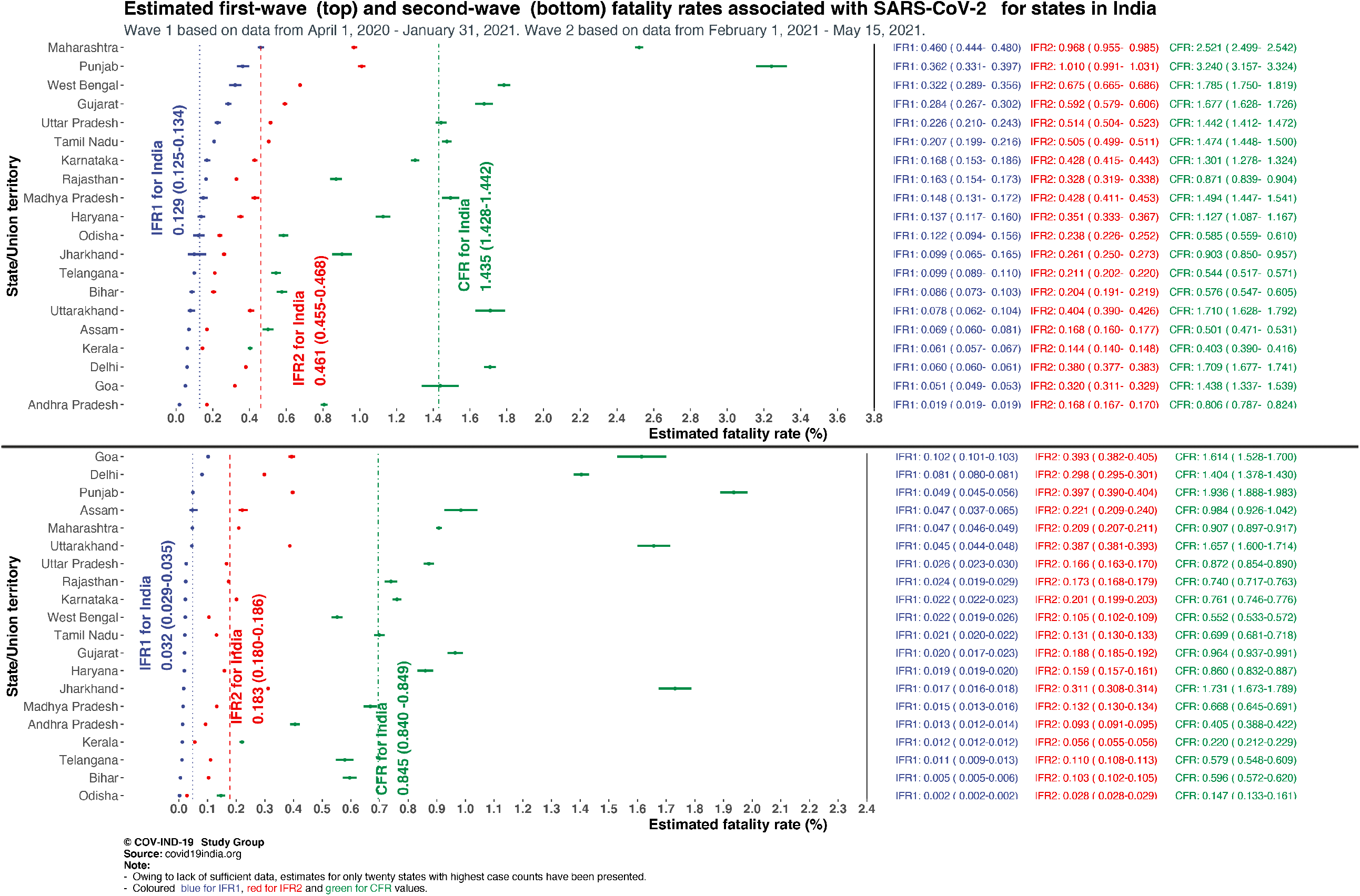
Forest plot of wave 1 and wave 2 infection fatality rates (IFR) and case fatality rates (CFR) associated with SARS-CoV-2 in various states in India. IFR1 is based on reported deaths whereas IFR2 estimates and includes the unreported deaths.

### Combining waves 1 and 2

The composite CFR as of May 15 stands at 1.1%. The estimate for total (reported + unreported) cumulative case count for waves 1 and 2 combined is ***491*.*73 (95% CrI 453*.*03 - 524*.*56)*** million, while the estimated number of total (reported + unreported) deaths is ***1216*.*35 (95% CrI 1154*.*21 - 1272*.*70)*** thousand. This leads to a combined IFR_1_ estimate of ***0*.*06%*** and IFR_2_ estimate of ***0*.*24%***. Detailed numerical estimates of underreporting factors across states for waves 1 and 2 are presented in ***Supplementary Tables T1, T2*** and ***T3*** and ***Supplementary Figures 2 and 3***.

## Discussion

Despite accounting for underreported deaths, the large number of asymptomatic/undetected infections (more than 90% by any calculation) indicate a lower IFR in India in comparison with other Western countries. A meta-analysis across the world places the pooled mean of IFRs at ***0*.*68% (95% CI: 0*.*53%-0*.*82%)***^***6***^, while another meta-analysis places the median at ***0*.*27%***^***7***^ ***(with a range of 0 - 1*.*63%)***. Seroprevalence surveys and epidemiologic models qualitatively agree on the estimated IFR for India for wave 1. Up to date serosurvey and excess death/mortality data are needed to validate wave 2 and combined estimates. The estimated number of total infections as of May 15 suggests roughly 36% of Indians have an active or past infection, a number that will need to be verified with synchronous sero-surveys.

The current reduction in fatality rates in wave 2 that we notice could be primarily due to two reasons, one is that we do not have the same length of follow-up period and complete data on the decay phase of wave 2 curve. The second could be the different age composition of the infected populations in the two waves; it has been reported that the younger population got infected in larger numbers in wave 2 and they have lower risk of COVID-19 mortality. A fraction of the older population (aged 65+ years) also got vaccinated during wave 2. However, this hypothesis about reduced fatality rates in wave 2 cannot be verified without more granular, age-sex stratified nationwide time-series data on case and death counts, which is currently unavailable.

## Limitations

We do not have a rigorous way to validate the extent of underreporting of deaths. An excess death calculation based on historical mortality data is infeasible at this point due to absence of all-cause-mortality data in the last three years from India. India has a very young population with only 6.4% in age group 65+ (compared to the US where this proportion is 16.5%) so a comparison of overall IFR between India and say the US is not fair, and only age-specific IFRs should be calculated and compared when more data become available. We do recognize that wave 2 information is appreciably incomplete, and the estimates will change as we have more complete information on deaths. For example, while our wave 2 analysis period ended on May 15, the highest daily number of deaths (4529 daily new deaths) were reported shortly after on May 18. Thus, our analysis presents an updated but incomplete picture of wave 2.

## Data Availability

All data are publicly available at covid19india.org. We used reported daily case, death and recovery counts for India and its states and union territories from April 1, 2020 to May 15, 2021. The statistical package SEIR-Fansy developed by the authors is available at covind19.org.

## List of abbreviations

CFR: Case Fatality Rate
CI: Confidence Interval
IFR: Infection Fatality Rate
RT-PCR: Real time reverse transcript polymerase chain reaction
SEIR: Susceptible-Exposed-Infected-Recovered
URF: Underreporting Factor

## Declarations

### Ethics approval and consent to participate

Not applicable. The analysis is based on publicly available completely de-identified aggregate counts. The study is exempt from IRB review as no patient participation or contact is involved.

### Consent to publish

All authors have consented for submission and potential publication in BMC Research Notes.

### Competing interests

There are no conflicts of interest perceived or declared by any of the authors.

### Funding

The research was supported by an internal pilot grant at the University of Michigan, awarded by the Michigan Institute of Data Science (MIDAS).

### Authors’ Contributions

Soumik Purkayastha created the initial draft, conducted the literature review and collaborated on the analysis. Ritoban Kundu and Ritwik Bhaduri created the R package SEIR-fansy and implemented the epidemiologic models. Dan Barker and Mike Kleinsasser collaborated on analysing data from the second wave. Debashree Ray helped create and modify the final draft and address issues raised by the review panel. Bhramar Mukherjee conceived the project, planned the analysis and wrote the draft of the paper. All authors read, reviewed, edited and approved the manuscript for submission.

## Acknowledgements

The authors are grateful for the computational resources available to them via the advanced research computing center at the University of Michigan.

## Notes

### Competing Interest Statement

The authors have declared no competing interest.

## References

[1]. COVIDIndia.org. COVID-19 Tracker Updates for India for State-wise and District-wise data. Published 2020. Accessed May 21, 2020. https://covidindia.org

[2]. Press Trust of India. Over 21% Of India’s Population May Have Had COVID-19, Shows Sero Survey. Published February 4, 2021. https://www.ndtv.com/india-news/over-21-of-indias-population-may-have-had-covid-19-shows-sero-survey-2363166

[3]. Wikipedia. Demographics of India: Structure of the population. Accessed February 20, 2021. https://en.wikipedia.org/wiki/Demographics_of_India#Structure_of_the_population

[4]. The Wire. COVID-19: Cremation, Burial Records Suggest Delhi’s Death Toll Is Over Twice the Official Figure. Published May 27, 2020. Accessed February 20, 2021. https://thewire.in/government/delhis-covid-19-deaths-data

[5]. Bhaduri R, Kundu R, Purkayastha S, Kleinsasser M, Beesley LJ, Mukherjee B. Extending the Susceptible-Exposed-Infected-Removed (SEIR) Model to Handle the High False Negative Rate and Symptom-Based Administration of COVID-19 Diagnostic Tests: SEIR-Fansy. Epidemiology; 2020. doi:10.1101/2020.09.24.20200238

[6]. Meyerowitz-Katz G, Merone L. A systematic review and meta-analysis of Published research data on COVID-19 infection fatality rates. Int J Infect Dis. 2020;101:138–148. doi:10.1016/j.ijid.2020.09.1464

[7]. Ioannidis JPA. Infection fatality rate of COVID-19 inferred from seroprevalence data. Bull World Health Organ. 2021;99(1):19–33F. doi:10.2471/BLT.20.265892

