## supplement for "Estimating the wave 1 and wave 2 infection fatality rates from SARS-CoV-2 in India"

***Supplementary material:*** ***Estimating the Wave 1 and Wave 2 infection fatality rate from SARS-CoV-2 in India***

***SEIR-fansy model***

***Introduction***

Here we are using the SEIR-fansy model^1^ and software package^2^ which uses a compartmental model accounting for false negative rates and preferential diagnostic testing for SARS-CoV-2 infections. The SEIR-fansy model can be represented by the compartmental model in Figure S1.

^1.^ Bhaduri R, Kundu R, Purkayastha S, Kleinsasser M, Beesley LJ, Mukherjee B. *Extending the Susceptible-Exposed-Infected-Removed (SEIR) model to handle the high false negative rate and symptom-based administration of Covid-19 diagnostic tests: SEIR-fansy.* medRxiv [Preprint]. 2020 Sep 25:2020.09.24.20200238. doi: 10.1101/2020.09.24.20200238. PMID: 32995829; PMCID: PMC7523173.

^2.^ Ritwik Bhaduri, Ritoban Kundu, Soumik Purkayastha, Lauren Beesley and Bhramar Mukherjee (2020). *SEIRfansy: Extended Susceptible-Exposed-Infected-Recovery Model.* R package version 1.1.0. https://CRAN.R-project.org/package=SEIRfansy

***Figure S1: Schematic diagram for the SEIR-fansy model with imperfect testing and misclassification.***

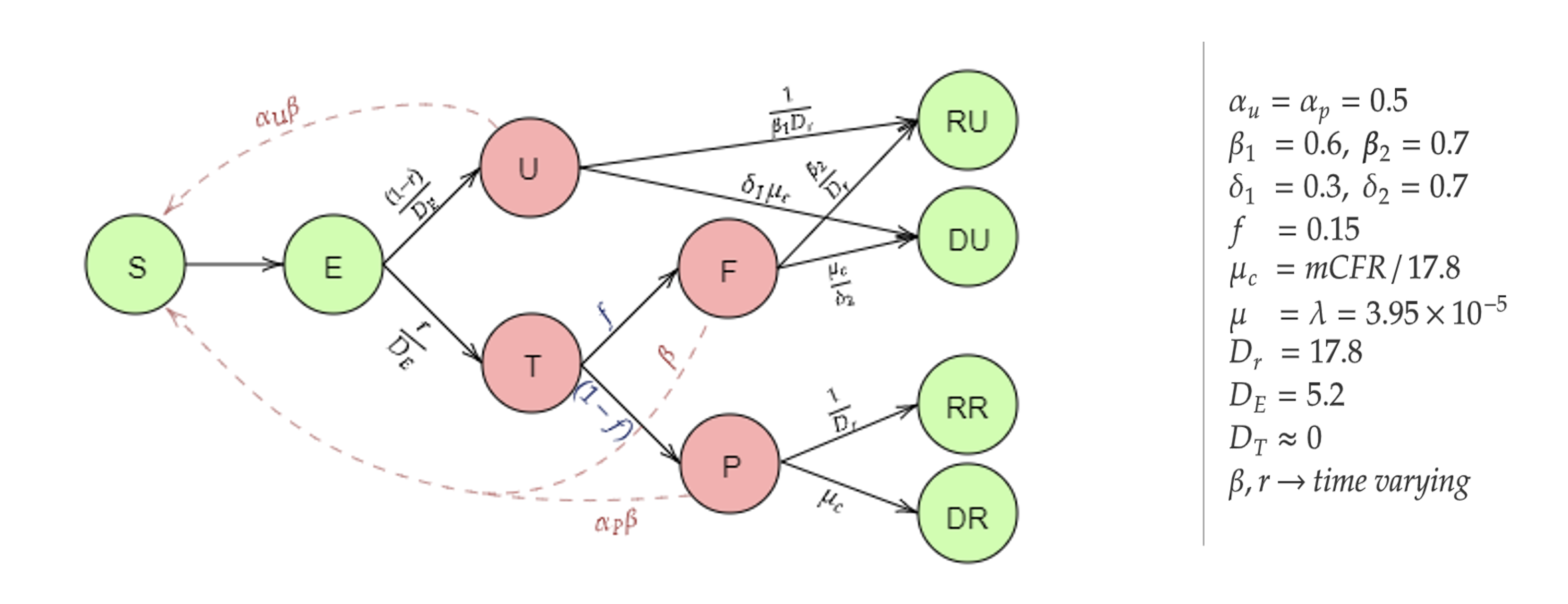

***Mathematical framework***

The following differential equations summarize the transmission dynamics being modeled.

$$\frac{\partial S}{\partial t}=-\beta\frac{S\left( t \right)}{N}\left( \alpha_{P}P\left( t \right)+\alpha_{U}U\left( t \right)+F\left( t \right) \right)+\lambda N-\mu S(t)$$

$$\frac{\partial E}{\partial t}=\beta\frac{S\left( t \right)}{N}\left( \alpha_{P}P\left( t \right)+\alpha_{U}U\left( t \right)+F\left( t \right) \right)-\frac{E\left( t \right)}{D_{e}}-\mu E\left( t \right)$$

$$\frac{\partial U}{\partial t}=\left( 1-r \right)\frac{E\left( t \right)}{D_{e}}-\frac{U\left( t \right)}{\beta_{1}D_{r}}-\delta_{1}\mu_{c}U\left( t \right)-\mu U\left( t \right)$$

$$\frac{\partial P}{\partial t}=\left( 1-f \right)r\frac{E\left( t \right)}{D_{e}}-\frac{P\left( t \right)}{D_{r}}-\mu_{c}P\left( t \right)-\mu P\left( t \right)$$

$$\frac{\partial F}{\partial t}=fr\frac{E\left( t \right)}{D_{e}}-\frac{\beta_{2}F\left( t \right)}{D_{r}}-\frac{\mu_{c}F\left( t \right)}{\delta_{2}}-\mu F(t)$$

$$\frac{\partial RU}{\partial t}=\frac{U\left( t \right)}{\beta_{1}D_{r}}+\frac{\beta_{2}F\left( t \right)}{D_{r}}-\mu RU\left( t \right)$$

$$\frac{\partial RR}{\partial t}=\frac{P\left( t \right)}{D_{r}}-\mu RR\left( t \right)$$

$$\frac{\partial DU}{\partial t}=\delta_{1}\mu_{c}U\left( t \right)+\frac{\mu_{c}F\left( t \right)}{\delta_{2}},$$

$$\frac{\partial DR}{\partial t}=\mu_{c}P\left( t \right)$$

Using the Next Generation Matrix Method (28), we have calculated the basic reproduction number

$$R_{0}=\frac{\beta S_{0}}{\mu D_{e}+1}\left( \frac{\alpha_{U}\left( 1-r \right)}{\frac{1}{\beta_{1}D_{r}}+\delta_{1}\mu_{c}+\mu}+\frac{\alpha_{P}r\left( 1-f \right)}{\frac{1}{D_{r}}+\mu_{c}+\mu}+\frac{rf}{\frac{\beta_{2}}{D_{r}}+\frac{\mu_{c}}{\delta_{2}}+\mu} \right)$$

where $S_{0}=\lambda/\mu=1$ since we have assumed that natural birth and death rates are equal within this short period of time. In this setting, both $\beta$ and $r$ are time-varying parameters which are estimated using the Metropolis-Hastings MCMC method. To estimate the parameters, we at first need to solve the differential equations, which is difficult to perform in this continuous-time setting. It is also worth noting that we do not require the values of the variables for each time point. Instead, we only need their values at discrete time steps, i.e., for each day. Thus, we approximate the above set of differential equations by a set of recurrence relations. For any compartment $X$, the instantaneous rate of change with respect to time $t$ (given by $\frac{\partial X}{\partial t}$) is approximated by the difference between the counts of that compartment on the $\left( t+1 \right)^{th}$ day and the $t^{th}$ day, that is $X\left( t+1 \right)-X(t)$. Starting with an initial value for each of the compartments on the Day 1 and using the discrete-time recurrence relations, we can then obtain the solutions of interest. Some examples of these discrete-time recurrence relations are presented below.

$$E\left( t+1 \right)-E\left( t \right)=\beta\frac{S\left( t \right)}{N}\left( \alpha_{P}P\left( t \right)+\alpha_{U}U\left( t \right)+F\left( t \right) \right)-\frac{E\left( t \right)}{D_{e}}-\mu E\left( t \right),$$

$$U\left( t+1 \right)-U\left( t \right)=\frac{\left( 1-r \right)E\left( t \right)}{D_{e}}-\frac{U\left( t \right)}{\beta_{1}D_{r}}-\delta_{1}\mu_{c} U\left( t \right)-\mu U\left( t \right),$$

$$P\left( t+1 \right)-P\left( t \right)=\frac{r\left( 1-f \right)E\left( t \right)}{D_{e}}-\frac{P\left( t \right)}{D_{r}}-\mu_{c}P\left( t \right)-\mu P\left( t \right),$$

$$F\left( t+1 \right)-F\left( t \right)=\frac{rfE\left( t \right)}{D_{e}}-\frac{\beta_{2}F\left( t \right)}{D_{r}}-\frac{\mu_{c} F\left( t \right)}{\delta_{2}}-\mu F\left( t \right).$$

The rest of the differential equations can each be similarly approximated by a discrete-time recurrence relation.

***Likelihood assumptions and estimation***

We use Bayesian estimation techniques and Markov chain Monte Carlo (MCMC) methods (namely, Metropolis-Hastings method with Gaussian proposal distribution) for estimating the parameters. First, we approximated the above set of differential equations using a discrete time approximation using daily differences. So, after we started with an initial value for each of the compartments on the day 1, using the discrete time recurrence relations we can find the counts for each of the compartments on the next days. To proceed with the MCMC-based estimation, we specify the likelihood explicitly. We assume (conditional on the parameters) the number of new confirmed cases on day $t$ depend only on the number of exposed individuals on the previous day. Specifically, we use multinomial modeling to incorporate the data on recovered and deceased cases as well. The joint conditional distribution is

$$P[P_{new}\left( t \right), R_{new}\left( t \right), D_{new}\left( t \right)|E\left( t-1 \right), P(t-1)]$$

$$=P[P_{new}\left( t \right)|E\left( t-1 \right), P(t-1)].P[ R_{new}\left( t \right), D_{new}\left( t \right)|E\left( t-1 \right), P(t-1)]$$

$=P[P_{new}\left( t \right)|E\left( t-1 \right)].P[ R_{new}\left( t \right), D_{new}\left( t \right)|P(t-1)]$

A multinomial distribution-like structure is then defined,

$$P_{new}\left( t \right)|E\left( t-1 \right) \sim Bin\left( E\left( t-1 \right),\frac{r\left( 1-f \right)}{D_{e}} \right)$$

$$R_{new}\left( t \right), D_{new}\left( t \right)|P\left( t-1 \right) \sim Mult\left( P\left( t-1 \right), \left( \frac{1}{D_{r}}, \mu_{c}, 1-\frac{1}{D_{r}}-\mu_{c} \right) \right)$$

*Note:* the expected values of $E(t-1)$ and $P(t-1)$ are obtained by solving the discrete time differential equations as described earlier.

***Prior assumptions and MCMC***

For the parameter $r$, we assume a $U(0,1)$ prior, while for $\beta$, we assume an improper non-informative flat prior with the set of positive real numbers as support. After specifying the likelihood and the prior distributions of the parameters, we draw samples from the posterior distribution of the parameters using the Metropolis-Hastings algorithm with a Gaussian proposal distribution. We run the algorithm for 200,000 iterations with a burn-in period of 100,000. Finally, the mean of the parameters in each of the iterations are obtained as the final estimates of $\beta$ and $r$ for the different time periods. To obtain confidence intervals of various estimates we predict the number of individuals in each compartment given a set of parameters which are drawn using MCMC. This is done for 100,000 iterations. Using these values, we obtain the 95% Bayesian Credible Intervals of the estimates (such as infection fatality rates and underreporting factors)

***Estimation of parameters of interest***

Our main parameters of interest here are Underreporting factors for cases and deaths and Infection Fatality rate. Underreporting factors (URF) for cases and deaths are defined as follows:

$$URF_{case} = \frac{Estimated Total Cumulative Infections}{Observed Cumulative Cases}$$

$$URF_{death} = \frac{Estimated Total Cumulative Deaths}{Observed Cumulative Deaths}$$

Here, total cumulative cases refers to all Cumulative cases including both reported and unreported cases. Similarly total cumulative deaths includes both reported and unreported deaths. Since we are unable to observe unreported cases or deaths we estimate total cumulative cases and deaths as follows:

1. Total Cumulative cases at time *t* = *P(t)+U(t)+F(t)+RR(t)+RU(t)+DR(t)+DU(t)*
2. Total Cumulative deaths at time *t* = *DR(t)+DU(t)*

Now, to estimate the true fatality rate of COVID-19, we calculate 2 different infection fatality rates:

$$IFR_{1}=\frac{Observed Cumulative Deaths}{Estimated Total Cumulative Infections}$$

$$IFR_{2}=\frac{Estimated Total Cumulative Deaths}{Estimated Total Cumulative Infections}$$

We also calculate the Case fatality rate which is defined as

$$CFR=\frac{Observed Cumulative Deaths}{Observed Cumulative Cases}$$

Now $Cumulative Deaths$follows a $Bin\left( Observed cumulative cases,CFR_{true} \right)$ distribution, with the estimate of $CFR_{true}$ given by CFR, making CFR is a binomial proportion . Let $\hat{p}=CFR$ and $n=Observed Cumulative Cases$ . So the asymptotic approximate 95% confidence interval is given by

($\hat{p}-1.96\sqrt{\frac{\hat{p}(1-\hat{p})}{n}}$,$\hat{p}+1.96\sqrt{\frac{\hat{p}(1-\hat{p})}{n}}$).

The estimates of infection fatality rates and underreporting factors are based on Bayesian credible intervals constructed from the exact posterior draws, as described before.

***Data source and results:***

The data has been sourced from [*covid19india.org*](https://www.covid19india.org/). We used daily case-recovery-death count data from April 1, 2020 to January 31, 2021 for wave 1 and from February 1, 2021 – May 15, 2021 for wave 2. The predicted number of reported and total cases and deaths for January 31, 2021 (wave 1) and May 15 (for wave 2 and waves 1 and 2 combined) are shown in Tables S1, S2, and S3 respectively.

The mean estimates and the 95% CrI’s of underreporting factors for cases and deaths on January 31, 2021 are shown in ***Figure S2.*** Relevant wave 2 values are presented in ***Figure S3.***

***Table T1: Summary of the different metrics for the states and the nation for wave 1, on 31st January, 2021***

| ***Place*** | ***Reported Cases (Observed)*** | ***Reported Cases (Predicted)*** | ***Total (reported + unreported) Cases (Predicted)*** | ***Reported Deaths (Observed)*** | ***Reported Deaths (Predicted)*** | ***Total (reported + unreported) Deaths (Predicted)*** | ***Cases per million*** | ***Deaths per million*** |
| --- | --- | --- | --- | --- | --- | --- | --- | --- |
| *Andaman and Nicobar* | *4990* | *4971* | *420209* | *62* | *62* | *1027* | *13111.53* | *162.91* |
| *Andhra Pradesh* | *887836* | *882030* | *38244770* | *7153* | *7125* | *64317* | *17908.19* | *144.28* |
| *Arunachal Pradesh* | *16828* | *16367* | *1177129* | *56* | *54* | *773* | *12161.36* | *40.47* |
| *Assam* | *217039* | *206824* | *1576369* | *1087* | *1050* | *2650* | *6955.14* | *34.83* |
| *Bihar* | *260719* | *265325* | *1752581* | *1501* | *1515* | *3566* | *2504.52* | *14.42* |
| *Chandigarh* | *20925* | *21064* | *370319* | *334* | *330* | *1456* | *19825.67* | *316.45* |
| *Chhattisgarh* | *305367* | *306365* | *3958345* | *3701* | *3573* | *12372* | *11953.99* | *144.88* |
| *Dadra and Nagar Haveli* | *3377* | *3353* | *322500* | *2* | *2* | *38* | *5765.12* | *3.41* |
| *Delhi* | *635096* | *634237* | *17949712* | *10853* | *10771* | *68212* | *37830.49* | *646.48* |
| *Goa* | *53409* | *53536* | *1516370* | *768* | *763* | *4854* | *36618* | *526.55* |
| *Gujarat* | *261539* | *239728* | *1546257* | *4386* | *3937* | *9160* | *4327.27* | *72.57* |
| *Haryana* | *267897* | *255093* | *2212182* | *3018* | *2849* | *7763* | *10567.32* | *119.05* |
| *Jammu and Kashmir* | *124506* | *120675* | *11786372* | *1936* | *1861* | *35906* | *10149.64* | *157.82* |
| *Jharkhand* | *118667* | *108448* | *1142000* | *1072* | *973* | *2982* | *3597.26* | *32.5* |
| *Karnataka* | *939387* | *931828* | *7298441* | *12224* | *12112* | *31189* | *15375.77* | *200.08* |
| *Kerala* | *929179* | *973432* | *6109388* | *3744* | *3821* | *8781* | *27814.68* | *112.08* |
| *Ladakh* | *9720* | *9875* | *195168* | *130* | *131* | *627* | *35474.45* | *474.45* |
| *Madhya Pradesh* | *255112* | *246254* | *2593075* | *3811* | *3617* | *11092* | *3512.64* | *52.47* |
| *Maharashtra* | *2026399* | *1939901* | *11106302* | *51081* | *48974* | *107479* | *18032.58* | *454.56* |
| *Manipur* | *29068* | *28426* | *787087* | *371* | *359* | *2239* | *11308.79* | *144.34* |
| *Meghalaya* | *13716* | *12742* | *496017* | *146* | *135* | *1126* | *4623.02* | *49.21* |
| *Mizoram* | *4372* | *4488* | *237273* | *9* | *9* | *98* | *3984.67* | *8.2* |
| *Nagaland* | *12057* | *11586* | *315789* | *82* | *83* | *506* | *6094* | *41.45* |
| *Odisha* | *335072* | *322600* | *1630327* | *1959* | *1882* | *3873* | *7982.8* | *46.67* |
| *Punjab* | *173276* | *176267* | *1553488* | *5615* | *5670* | *15692* | *6245.68* | *202.39* |
| *Rajasthan* | *317491* | *295702* | *1696333* | *2766* | *2552* | *5569* | *4631.63* | *40.35* |
| *Sikkim* | *6104* | *6136* | *177128* | *133* | *135* | *877* | *9997.1* | *217.83* |
| *Tamil Nadu* | *838340* | *842658* | *5966633* | *12356* | *12378* | *30119* | *11619.88* | *171.26* |
| *Telangana* | *293959* | *287679* | *1625763* | *1599* | *1560* | *3426* | *8397.95* | *45.68* |
| *Tripura* | *33347* | *32051* | *801142* | *388* | *374* | *2131* | *9076.69* | *105.61* |
| *Uttar Pradesh* | *600299* | *584173* | *3838576* | *8658* | *8418* | *19731* | *3004.31* | *43.33* |
| *Uttarakhand* | *96129* | *94366* | *2147503* | *1644* | *1609* | *8655* | *9530.66* | *162.99* |
| *West Bengal* | *569998* | *548980* | *3171287* | *10173* | *9735* | *21392* | *6244.77* | *111.45* |
| *India* | *10758629* | *10512888* | *119510413* | *154428* | *149478* | *550380* | *8022.84* | *115.16* |

***Table T2: Summary of the different metrics for the states and the nation for wave 2, on 15th May, 2021***

| ***Place*** | ***Reported Cases (Observed)*** | ***Reported Cases (Predicted)*** | ***Total (reported + unreported) Cases (Predicted)*** | ***Reported Deaths (Observed)*** | ***Reported Deaths (Predicted)*** | ***Total (reported + unreported) Deaths (Predicted)*** | ***Cases per million*** | ***Deaths per million*** |
| --- | --- | --- | --- | --- | --- | --- | --- | --- |
| *Andaman and Nicobar* | *1609* | *1787* | *19673* | *26* | *20* | *69* | *4227.75* | *68.32* |
| *Andhra Pradesh* | *547591* | *590628* | *17051318* | *2218* | *1995* | *15836* | *11045.24* | *44.74* |
| *Arunachal Pradesh* | *4975* | *5781* | *34678* | *25* | *22* | *54* | *3595.36* | *18.07* |
| *Assam* | *111475* | *135174* | *2360088* | *1096* | *1011* | *5197* | *3572.28* | *35.12* |
| *Bihar* | *391115* | *454292* | *43056947* | *2329* | *1958* | *44517* | *3757.13* | *22.37* |
| *Chandigarh* | *34410* | *36951* | *637191* | *301* | *271* | *1294* | *32602.21* | *285.19* |
| *Chhattisgarh* | *606788* | *658814* | *20557122* | *8028* | *7241* | *56275* | *23753.51* | *314.27* |
| *Dadra and Nagar Haveli* | *6138* | *7457* | *68901* | *2* | *2* | *6* | *10478.62* | *3.41* |
| *Delhi* | *758650* | *872338* | *13190453* | *10650* | *8986* | *39319* | *45190.18* | *634.38* |
| *Goa* | *82387* | *88631* | *1305218* | *1331* | *1217* | *5135* | *56485.74* | *912.55* |
| *Gujarat* | *490781* | *546922* | *24250682* | *4733* | *4288* | *45594* | *8120.18* | *78.31* |
| *Haryana* | *426438* | *480984* | *19159948* | *3663* | *3034* | *30441* | *16821.04* | *144.49* |
| *Jammu and Kashmir* | *120058* | *121462* | *8731279* | *1213* | *1139* | *19689* | *9787.05* | *98.88* |
| *Jharkhand* | *196767* | *224000* | *20256985* | *3406* | *2939* | *63052* | *5964.78* | *103.25* |
| *Karnataka* | *1263687* | *1264630* | *42843833* | *9617* | *9900* | *86103* | *20683.87* | *157.41* |
| *Kerala* | *1215330* | *1264428* | *22365420* | *2668* | *2559* | *12414* | *36380.52* | *79.87* |
| *Ladakh* | *6728* | *7545* | *87363* | *35* | *33* | *119* | *24554.74* | *127.74* |
| *Madhya Pradesh* | *476122* | *509966* | *21838965* | *3180* | *2815* | *28811* | *6555.73* | *43.79* |
| *Maharashtra* | *3350105* | *3565542* | *64446203* | *30377* | *27553* | *134626* | *29812.01* | *270.32* |
| *Manipur* | *10651* | *10736* | *31428* | *207* | *209* | *390* | *4143.73* | *80.53* |
| *Meghalaya* | *9568* | *10401* | *68564* | *174* | *157* | *414* | *3224.93* | *58.65* |
| *Mizoram* | *4307* | *4827* | *35364* | *15* | *15* | *41* | *3925.43* | *13.67* |
| *Nagaland* | *5977* | *5805* | *12841* | *121* | *202* | *357* | *3020.97* | *61.16* |
| *Odisha* | *277073* | *307713* | *18117660* | *407* | *353* | *5127* | *6601.03* | *9.7* |
| *Punjab* | *324235* | *340007* | *12891875* | *6279* | *5527* | *51111* | *11686.95* | *226.32* |
| *Rajasthan* | *542082* | *570565* | *17180159* | *4011* | *3772* | *29718* | *7908.01* | *58.51* |
| *Sikkim* | *5319* | *5374* | *35209* | *72* | *95* | *259* | *8711.43* | *117.92* |
| *Tamil Nadu* | *759374* | *838877* | *25501756* | *5307* | *4315* | *33460* | *10525.37* | *73.56* |
| *Telangana* | *234236* | *258389* | *12601245* | *1354* | *1179* | *13909* | *6691.75* | *38.68* |
| *Tripura* | *7465* | *6780* | *29447* | *40* | *42* | *92* | *2031.89* | *10.89* |
| *Uttar Pradesh* | *1019175* | *1182153* | *34787622* | *8884* | *7691* | *57859* | *5100.66* | *44.46* |
| *Uttarakhand* | *191106* | *204540* | *6964680* | *3163* | *3139* | *26952* | *18947.1* | *313.59* |
| *West Bengal* | *563253* | *664641* | *14075639* | *3105* | *2571* | *14738* | *6170.87* | *34.02* |
| *India* | *14197732* | *15114765* | *379550762* | *119895* | *105254* | *692385* | *10587.42* | *89.41* |

***Table T3: Summary of the different metrics for the states and the nation for waves 1 and 2 combined, on 15th May, 2021.***

| ***Place*** | ***Reported Cases (Observed)*** | ***Reported Cases (Predicted)*** | ***Total (reported + unreported) Cases (Predicted)*** | ***Reported Deaths (Observed)*** | ***Reported Deaths (Predicted)*** | ***Total (reported + unreported) Deaths (Predicted)*** | ***Cases per million*** | ***Deaths per million*** |
| --- | --- | --- | --- | --- | --- | --- | --- | --- |
| *Andaman and Nicobar* | *1609* | *1787* | *19673* | *26* | *20* | *69* | *4227.75* | *68.32* |
| *Andhra Pradesh* | *547591* | *590628* | *17051318* | *2218* | *1995* | *15836* | *11045.24* | *44.74* |
| *Arunachal Pradesh* | *4975* | *5781* | *34678* | *25* | *22* | *54* | *3595.36* | *18.07* |
| *Assam* | *111475* | *135174* | *2360088* | *1096* | *1011* | *5197* | *3572.28* | *35.12* |
| *Bihar* | *391115* | *454292* | *43056947* | *2329* | *1958* | *44517* | *3757.13* | *22.37* |
| *Chandigarh* | *34410* | *36951* | *637191* | *301* | *271* | *1294* | *32602.21* | *285.19* |
| *Chhattisgarh* | *606788* | *658814* | *20557122* | *8028* | *7241* | *56275* | *23753.51* | *314.27* |
| *Dadra and Nagar Haveli* | *6138* | *7457* | *68901* | *2* | *2* | *6* | *10478.62* | *3.41* |
| *Delhi* | *758650* | *872338* | *13190453* | *10650* | *8986* | *39319* | *45190.18* | *634.38* |
| *Goa* | *82387* | *88631* | *1305218* | *1331* | *1217* | *5135* | *56485.74* | *912.55* |
| *Gujarat* | *490781* | *546922* | *24250682* | *4733* | *4288* | *45594* | *8120.18* | *78.31* |
| *Haryana* | *426438* | *480984* | *19159948* | *3663* | *3034* | *30441* | *16821.04* | *144.49* |
| *Jammu and Kashmir* | *120058* | *121462* | *8731279* | *1213* | *1139* | *19689* | *9787.05* | *98.88* |
| *Jharkhand* | *196767* | *224000* | *20256985* | *3406* | *2939* | *63052* | *5964.78* | *103.25* |
| *Karnataka* | *1263687* | *1264630* | *42843833* | *9617* | *9900* | *86103* | *20683.87* | *157.41* |
| *Kerala* | *1215330* | *1264428* | *22365420* | *2668* | *2559* | *12414* | *36380.52* | *79.87* |
| *Ladakh* | *6728* | *7545* | *87363* | *35* | *33* | *119* | *24554.74* | *127.74* |
| *Madhya Pradesh* | *476122* | *509966* | *21838965* | *3180* | *2815* | *28811* | *6555.73* | *43.79* |
| *Maharashtra* | *3350105* | *3565542* | *64446203* | *30377* | *27553* | *134626* | *29812.01* | *270.32* |
| *Manipur* | *10651* | *10736* | *31428* | *207* | *209* | *390* | *4143.73* | *80.53* |
| *Meghalaya* | *9568* | *10401* | *68564* | *174* | *157* | *414* | *3224.93* | *58.65* |
| *Mizoram* | *4307* | *4827* | *35364* | *15* | *15* | *41* | *3925.43* | *13.67* |
| *Nagaland* | *5977* | *5805* | *12841* | *121* | *202* | *357* | *3020.97* | *61.16* |
| *Odisha* | *277073* | *307713* | *18117660* | *407* | *353* | *5127* | *6601.03* | *9.7* |
| *Punjab* | *324235* | *340007* | *12891875* | *6279* | *5527* | *51111* | *11686.95* | *226.32* |
| *Rajasthan* | *542082* | *570565* | *17180159* | *4011* | *3772* | *29718* | *7908.01* | *58.51* |
| *Sikkim* | *5319* | *5374* | *35209* | *72* | *95* | *259* | *8711.43* | *117.92* |
| *Tamil Nadu* | *759374* | *838877* | *25501756* | *5307* | *4315* | *33460* | *10525.37* | *73.56* |
| *Telangana* | *234236* | *258389* | *12601245* | *1354* | *1179* | *13909* | *6691.75* | *38.68* |
| *Tripura* | *7465* | *6780* | *29447* | *40* | *42* | *92* | *2031.89* | *10.89* |
| *Uttar Pradesh* | *1019175* | *1182153* | *34787622* | *8884* | *7691* | *57859* | *5100.66* | *44.46* |
| *Uttarakhand* | *191106* | *204540* | *6964680* | *3163* | *3139* | *26952* | *18947.1* | *313.59* |
| *West Bengal* | *563253* | *664641* | *14075639* | *3105* | *2571* | *14738* | *6170.87* | *34.02* |
| *India* | *14197732* | *15114765* | *379550762* | *119895* | *105254* | *692385* | *10587.42* | *89.41* |

***Table T4:*** ***Parameter values and descriptions for the SEIRfansy model.***

| **Parameter** | **Value** | **Description** |
| --- | --- | --- |
|  | *Time-varying* | Rate of infectious transmission by infected, tested individuals with false negative results. |
| p | 0.5 | Ratio of rate of spread of infection by tested positive patients to that by false negatives. p < 1 represents the scenario where individuals who test positive are infecting susceptible individuals are a lower rate than infected individuals with false negative test results. |
| u | 0.5 | Scaling factor for the rate of spread of infection by untested individuals. u is assumed to be < 1 as U mostly consists of asymptomatic or mildly symptomatic cases who are known to spread the disease at a much lower rate than those with higher levels of symptoms. |
| De | 5.2 | Incubation period (in days). |
| Dr | 17.8 | Means number of days until recovery for infected individuals. |
| Dt | 0 | Mean number of days for the test result to come after a person is tested. Under the assumption of instantaneous test results, this is taken to be zero. |
| c | 0.0562 | Death rate attributable to COVID-19 which is equivalent to inverse of the average number of days for death starting from the onset of disease times the probability of death of an infected individual. |
| λ, μ | 3.95x10^-5^ | Natural birth and death rates (assumed to be equal). |
| r | *Time-varying* | Probability of being tested for infectious individuals. |
| f | 0.15 | Probability of a false negative RT-PCR diagnostic test result. |
| 1, 12 | 0.6 (1)  0.7 (2) | Scaling factors for rate of recovery for undetected and false negative individuals respectively. Both 1 and 2 are assumed to be less than 1. It is assumed that the recovery rate is slower than the detected ones for the False Negative ones because they are not getting any hospital treatments. The condition of Untested individuals is not so severe as they consist of mostly asymptomatic people. So, they are assumed to recover faster than the Current Positive Ones. |
| 1, 12 | 0.3 (1)  0.7 (2) | Scaling factors for death rate for undetected and false negative individuals respectively. Both 1 and 2 are assumed to be less than 1. Same as before, the death rate for False Negative ones are assumed to be higher than the Current detected Positive as they are not receiving proper treatment. While, for the Untested ones, the death rate is taken to be lesser because they are mostly asymptomatic. So, their probability of dying is much less. |

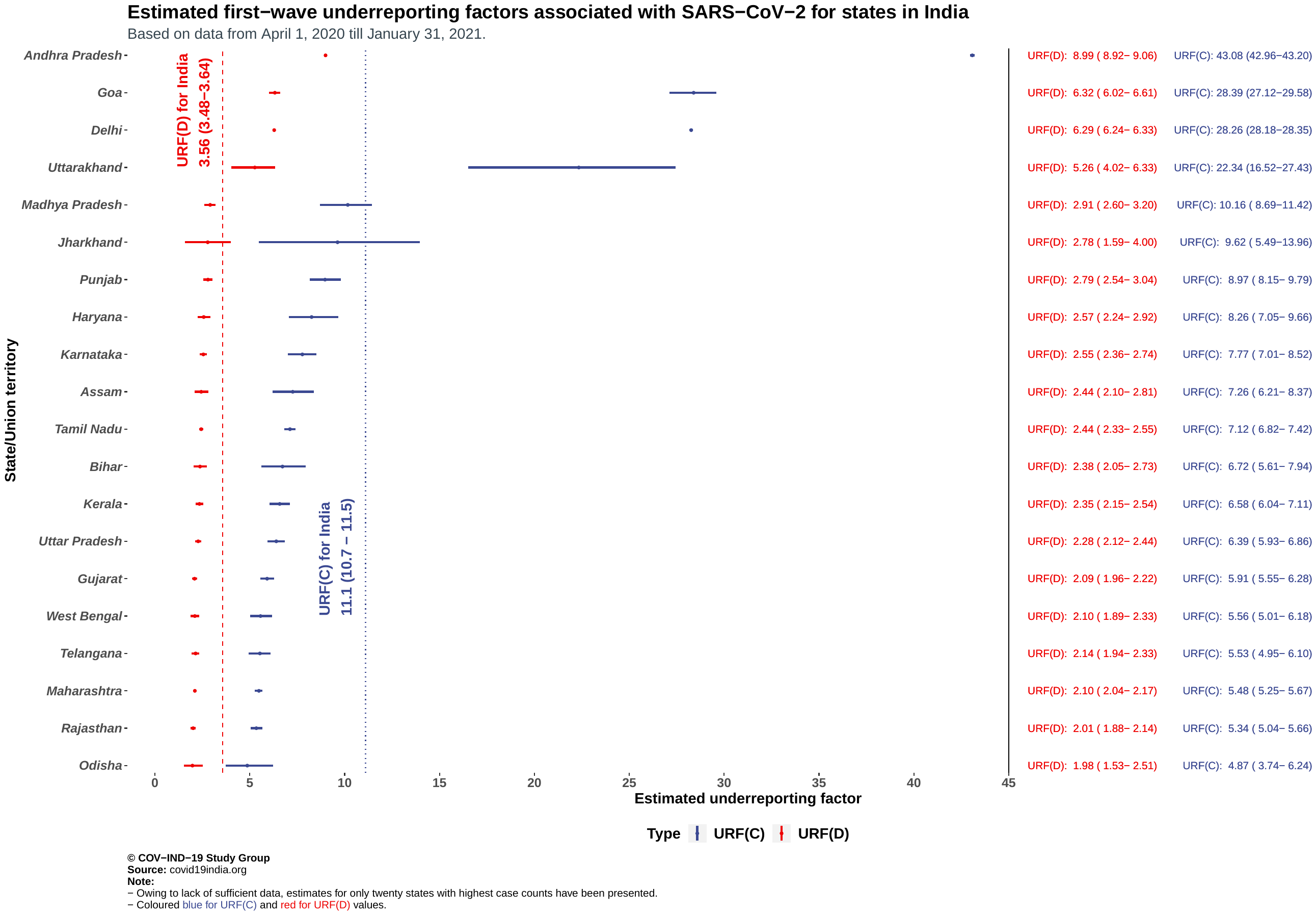

**Supplementary Figure 2: Estimated first wave underreporting factors for cases and deaths associated with SARS-CoV-2 for states in India.**

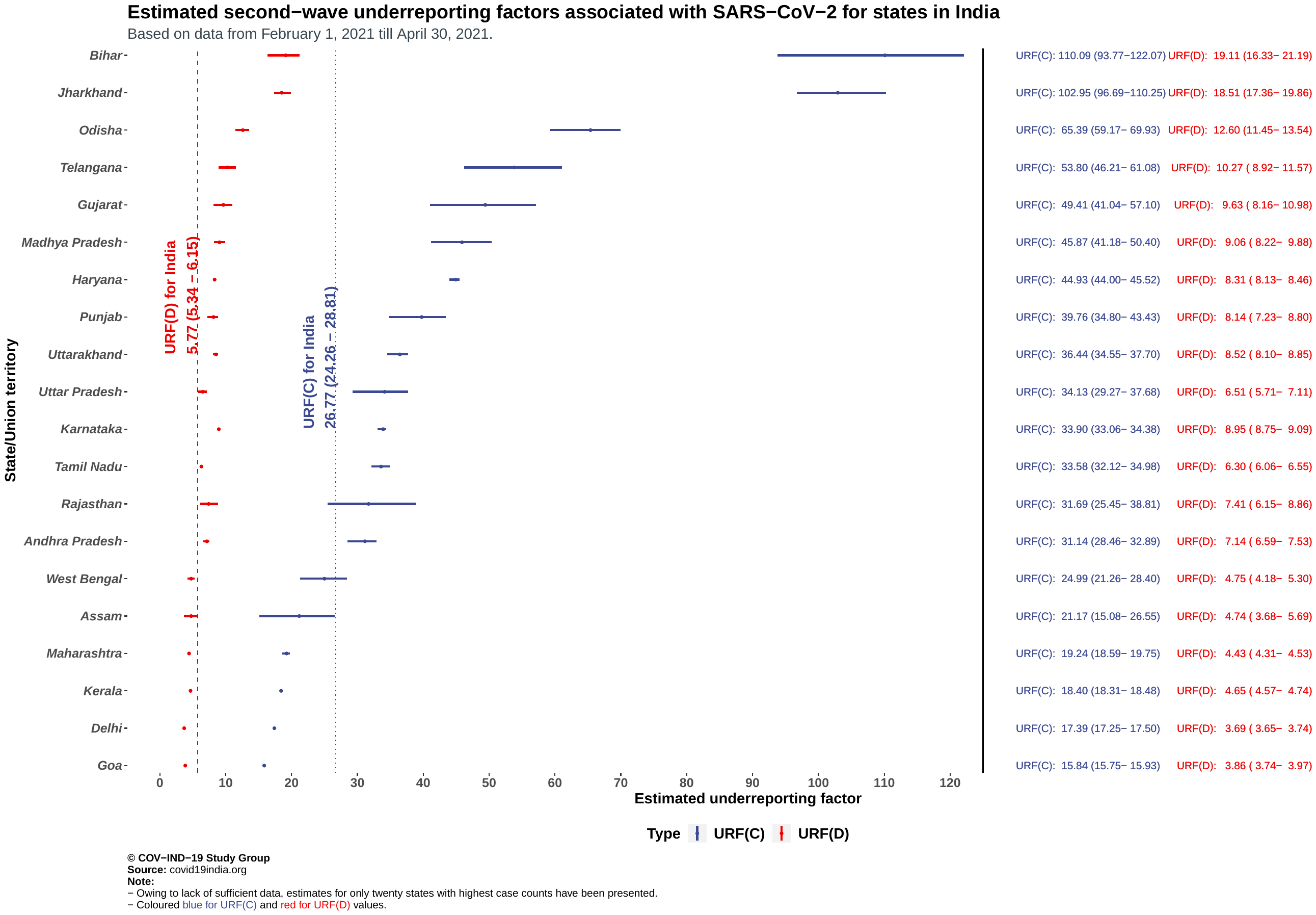

**Supplementary Figure 3: Estimated first wave underreporting factors for cases and deaths associated with SARS-CoV-2 for states in India.**
